# Effects of Systemic Lupus Erythematosus on the Brain: A Systematic Review of Structural MRI Findings and their Relationships with Cognitive Dysfunction

**DOI:** 10.1101/2024.03.12.24304183

**Authors:** Diana Valdés Cabrera, Tala El Tal, Ibrahim Mohamed, Santiago Eduardo Arciniegas, Stephanie Fevrier, Justine Ledochowski, Andrea Knight

## Abstract

**Background:** Cognitive dysfunction (CD) is highly prevalent in systemic lupus erythematosus (SLE), yet the underlying mechanisms are poorly understood. Neuroimaging utilizing advanced magnetic resonance imaging (MRI) metrics may yield mechanistic insights. We conducted a systematic review of neuroimaging studies to investigate the relationship between structural and diffusion MRI metrics and CD in SLE.

**Methods:** We systematically searched several databases between January 2000 and October 2023 according to Preferred Reporting Items for Systematic Reviews and Meta-Analyses (PRISMA) guidelines. Retrospective, and prospective studies were screened for search criteria keywords (including structural or diffusion MRI, cognitive function, and SLE) to identify peer-reviewed articles reporting advanced structural MRI metrics and evaluating CD in human patients with SLE.

**Results:** Eighteen studies (8 structural MRI, 9 diffusion MRI, and 1 with both modalities) were included; sample sizes ranged from 11 to 120 participants with SLE. Neurocognitive assessments and neuroimaging techniques, parameters, and processing differed across articles. The most frequently affected cognitive domains were memory, psychomotor speed, and attention; while abnormal structural and/or diffusion MRI metrics were found more consistently in the hippocampus, corpus callosum, and frontal cortex of patients with SLE, with and without clinically diagnosed CNS involvement.

**Conclusion:** Advanced structural MRI analysis can identify total and regional brain abnormalities associated with CD in patients with SLE, with potential to enhance clinical assessment. Future collaborative, longitudinal studies of neuroimaging in SLE are needed to better characterize CD, with focus on harmonized neurocognitive assessments, neuroimaging acquisitions and post-processing analyses, and improved clinical characterization of SLE cohorts.

## Introduction

Neuropsychiatric systemic lupus erythematosus (NPSLE) remains one of the most challenging manifestations of SLE due to the broad spectrum of symptoms (up to 19 clinical syndromes affecting either the central – CNS or the peripheral nervous system) and limited understanding of the underlying disease neurobiology^1^. These syndromes can be categorized into focal or diffuse, with the latter manifestations showing highest prevalence, mainly as cognitive dysfunction (CD, ∼80%)^2,3^ and mood disturbances (∼65%)^1,3^. In addition, neuropsychiatric involvement occurs more frequently in childhood-onset SLE (cSLE, up to 95%) than in adult-onset SLE (aSLE, 11-81%), and it strikes during a critical period of neurodevelopment that may potentially lead to irreversible negative impact in cognitive function^4^.

CD in SLE has been defined by the American College of Rheumatology (ACR) as a significant deficit in one or several of the following domains: attention, reasoning, executive skills, memory, visual-spatial processing, language, and psychomotor speed^5^, with attention and memory amongst the most regularly affected cognitive domains^6,7^. CD due to NPSLE could be a consequence of several pathologic mechanisms related to vascular involvement, blood-brain barrier breach and cell-mediated inflammation^8^. Yet, NPSLE pathogenesis is still poorly understood, making diagnosis and monitoring particularly challenging. CD may also be due to other factors and therefore difficult to attribute to NPSLE^7,8^.

Magnetic resonance imaging (MRI) is the gold standard neuroimaging tool to diagnose and monitor NPSLE^1,9^. However, conventional structural MRI abnormalities, such as white matter (WM) hyperintensities, gross brain tissue atrophy and ventricular enlargement in response to this atrophy are not always observed in patients with NPSLE^10^. In contrast, more advanced post-processing techniques enable the quantification of structural brain metrics beyond total tissue volumes from standard T1-weighted MRI, such as regional volumes, surface area, and cortical grey matter (GM) thickness^11,12^. These metrics can be semi-automatically calculated from human brain atlases and can reveal subtle NPSLE-related brain abnormalities (e.g., frontal cortex atrophy), not apparent with more conventional clinical tools^10–12^.

In addition to these post-processing methods, other less conventional structural MRI sequences, specifically diffusion MRI, have become a subject of interest in current NPSLE clinical research^10,13^. Diffusion MRI measures the random motion of water molecules (diffusion coefficient D) while they interact with tissue boundaries, cell membranes and other biological barriers, yielding structural metrics linked to axonal loss, inflammation, and demyelination, particularly in the WM^14^; thus, it can probe the status of brain tissue microstructure in relationship to neurological symptoms. Common metrics include the mean (MD), axial (AD), and radial (RD) diffusivities, which respectively characterize water diffusion in bulk, parallel, and perpendicular to a WM tract, and fractional anisotropy (FA), which quantifies the degree of diffusion anisotropy or directionality in a voxel^15,16^. Additionally, diffusion weighted imaging (DWI) can be utilized to weight the strength of WM connections by quantifying properties of brain-wide structural networks (e.g., node strength, density)^17^, and to evaluate intra-voxel incoherent-motion (IVIM), where the microcirculation of blood-water in the capillary network (D*), and tissue D and perfusion can be estimated^18^.

An increasing number of studies are utilizing advanced MRI to investigate NPSLE^10^. Altered tissue microstructure has been reported in several brain regions of patients with and without NPSLE diagnosis when compared to healthy controls^11,19–21^, and they have correlated with higher CD^20^. Overall, the presence of these associations even in the absence of clinical NPSLE diagnosis, suggest that brain involvement could be underdetected in SLE. However, existing advanced neuroimaging research in SLE has been limited in generalizability and interpretation due to small cohorts, and often incomplete characterization of clinical features. In response to these knowledge gaps, we conducted a systematic review to evaluate (i) the effect of SLE on brain structure, (ii) the neuroimaging correlates of CD in SLE, and (iii) potential disease-related contributors including but not limited to disease activity, duration and glucocorticoid exposure. A better understanding of these associations will help inform attribution of CD to SLE, characterization of domain specific CD trajectories, and possible new therapeutic strategies for protection of cognitive function in children and adults diagnosed with SLE.

## Methods

### Search Strategy

This systematic review was conducted in agreement with the Preferred Reporting Items for Systematic Reviews and Meta-Analyses (PRISMA) guidelines^22^. The search was performed in PubMed, MEDLINE, Embase, Web of Science, and Cochrane databases, and it was aided by the web-based literature review manager Covidence. It included the following terms: ‘systemic lupus erythematosus (SLE)’, OR ‘neuropsychiatric lupus (NPSLE)’, OR ‘central nervous system (CNS) lupus’, OR ‘antiphospholipid syndrome SLE’; AND ‘magnetic resonance imaging (MRI, structural MRI)’; OR ‘diffusion MRI (diffusion tensor imaging – DTI, DWI)’.

### Inclusion and Exclusion Criteria

#### Inclusion Criteria

(i) Peer-reviewed articles, limited to human research, and published between January 2000 and October 2023, including observational, case series, cross-sectional, longitudinal, retrospective, or prospective study designs of SLE populations; (ii) neuroimaging studies that utilized structural (T1-weighted) and/or diffusion MRI; (iii) evaluation of cognitive function/performance in SLE.

#### Exclusion Criteria

(i) Reviews, meta-analyses, and manuscripts that do not refer to MRI data directly collected from SLE cohorts; (ii) studies solely reporting conventional T1-weighted MRI metrics, such as total brain volumes (and not regional volumes), lateral ventricles volume, and/or WM hyperintensities numbers/volumes.

### Identification of Eligible Studies

Title and abstracts were reviewed for eligibility by six team members (DVC, TE, IM, SA, SF, JL). A full text review of potentially eligible articles according to inclusion and exclusion criteria was undertaken independently by these members, and afterwards final articles were selected by consensus.

### Data Extraction

Information extracted from studies included: main publication details (first author, year, country), study design (cross-sectional, longitudinal), cohort demographics (sample size, age, sex, ethnicity), clinical variables (NPSLE clinical diagnosis, disease duration, SLE Disease Activity Index – SLEDAI scores, Systemic Lupus International Collaborating Clinic Damage Index – SDI scores, glucocorticoid use), CD assessments (cognitive domains, neuropsychological tests, CD definitions), MRI technical details (magnetic fields, voxel sizes, b-values, diffusion directions), protocol type (structural or diffusion MRI sequences), and structural (total GM, WM, and regional volumes; cortical thickness) and diffusion MRI metrics (FA, MD, AD, RD). Two reviewers (DVC and TE) individually extracted data from the included articles regarding associations between atypical structural and/or diffusion brain MRI metrics and CD in SLE. Associations between brain MRI abnormalities and other clinical variables were summarized when available.

## Results

A total of 18 articles that evaluated the effect of SLE on brain structure and their links with CD were included in this review (Figure 1)^12,19–21,23–36^. From these studies, eight utilized T1-weighted MRI^12,28,29,31–33,35,36^, nine focused on diffusion MRI^19–21,24–27,30,34^, and one reported brain metrics from both modalities^23^. Most studies (15/18) were cross-sectional in design, while three studies assessed longitudinal changes in brain structure in their SLE cohorts at two different time points^25,31,35^.

**Figure 1:**
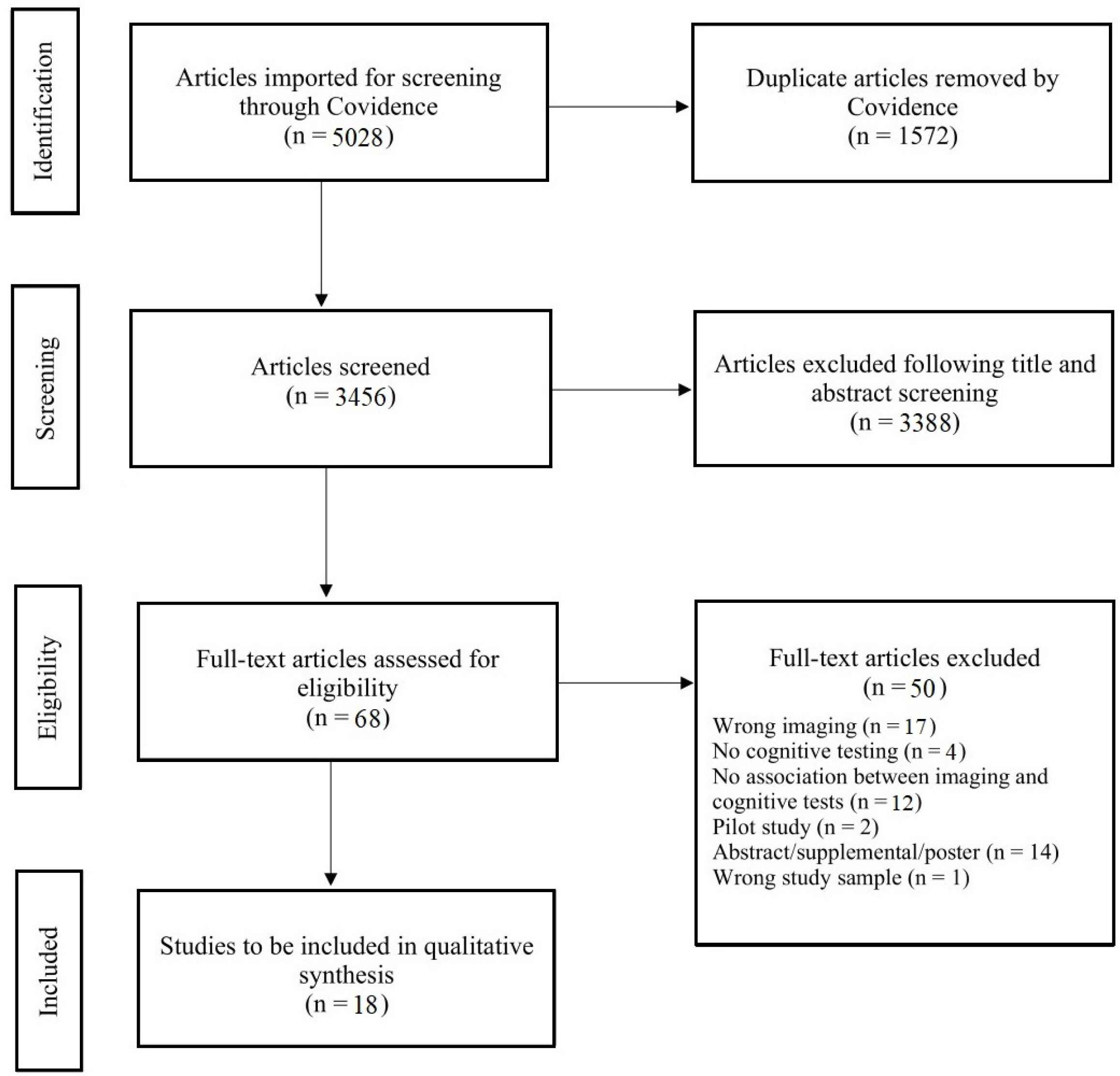
Preferred Reporting Items for Systematic Reviews and Meta-Analyses (PRISMA) flow diagram utilized to screen existing MRI studies in SLE.

### Sample Sizes, Demographics, and Clinical Features

An overview of demographic and clinical characteristics of the SLE cohorts is presented in Table 1. SLE sample sizes ranged from 11 to 120 with a mean of 50 participants, and in all cohorts at least 80% were females (17/18 papers reported biological sex as part of patient’s demographics) which is in line with the highest prevalence of SLE in women^37^. Three manuscripts reported information regarding race/ethnicity, and in two of them over half of the patients were African-Americans (6/11 – 55%, and 14/20 – 70% of their respective cohorts^25,27^). Cohort ages ranged 14.7-48.9 years on average with a pooled mean of 35.5 years; 16/18 studies examined adult patients with SLE, 2/18 included patients with cSLE^27,28^, and one compared patients with aSLE versus cSLE^26^. Healthy controls with age and sex distributions comparable to their respective SLE cohorts were utilized for group comparisons in 16/18 studies. Only three studies longitudinally evaluated 56%, 65%, and 100% of their SLE cohorts at follow-up time points that ranged 12-19 months from the date of their respective baseline MRI scans^25,31,35^.

**Table 1:** Demographic and Clinical Characteristics of SLE Cohorts (Least to Most Recent Publication Date).

| **First Author, Year, Country (Study Design)** | **Group: Size N, Age (years): Mean (SD) (or Range, 95% CI), Sex: % Females - F** | **Disease Duration (years): Mean (SD) (or Median, IQR)** | **Disease Activity (SLEDAI, SLAM) & Damage (SDI): Mean (SD)** | **Steroid Use: % Patients (cr - current, cm - cumulative)** |
| --- | --- | --- | --- | --- |
| Appenzeller, 2006, Brazil<br>(Longitudinal) | SLE: N=107, 32.2 (11.2) years, 93.5% F<br>Follow-up (19 months): N=60<br>HC: N=40, 31.8 (10.2) years, 90% F | 5.4 (4.0) | SLEDAI: 14.0 (5.9), SDI: 2.3 (1.8)<br>Follow-up: SLEDAI: 12.4 (3.8), SDI: 3.1 (1.7) | 100% - 107 SLE (cm) |
| Appenzeller, 2007, Brazil<br>(Longitudinal) | SLE: N=75, 32.3 (12.5) years, 93% F<br>Follow-up ($\geq 12$ months): N=75<br>HC: N=44, 33.8 (13.7) years, 91% F | 5.4 (4.5) | SLEDAI: 15.3 (8.3), SDI: 2.0 (2.1)<br>Follow-up: NR | 100% - 75 SLE (cm) |
| Jung, 2012, USA<br>(Cross-sectional) | Non-NPSLE: N=15, 37.5 (Range=18-59) years, 100% F<br>NPSLE: N=16, 38.1 (Range=18-59) years, 100% F<br>HC: N=18, 32.2 years, 100% F | NR | NR | 87% - 13/15 Non-NPSLE (cr)<br>94% - 15/16 NPSLE (cr) |
| Gitelman, 2013, USA<br>(Cross-sectional) | cSLE w/o CD: N=14, 14.7 (2.1) years, 79% F<br>cSLE w/ CD: N=8, 15.2 (1.8) years, 88% F<br>HC: N=19, 14.3 (2.2) years, 79% F | 2.5 (2.2)<br>2.0 (1.8) | SLEDAI: 4.1 (3.0), SDI: 0.4 (0.7)<br>SLEDAI: 8.8 (7.3), SDI: 0.6 (1.1) | 36% - 5/14 w/o CD (cr & cm)<br>62.5% - 5/8 w/ CD (cr & cm) |
| Cesar, 2015, USA<br>(Cross-sectional) | NPSLE: N=23, 48.9 (12.5) years, 95.7% F<br>MS: N=30, 43.8 (8.6) years, 76.5% F<br>HC: N=43, 44.7 (9.8) years, 86% F | 15.0 (9.6) | SLAM: 9.9 (4.9)<br>(Mean to moderate disease severity for NPSLE & MS) | NR |
| Bizzo, 2016, Brazil<br>(Cross-sectional) | SLE w/o EM deficit: N=34, 45.9 (11.4) years<br>SLE w/ EM deficit: N=17, 43.9 (10.3) years<br>HC: N=34, 45.7 (9.7) years | > 0.5 | SDI: 1.8 (1.6)<br>SDI: 0.9 (1.2) | NR |
| Bódi, 2017, Hungary<br>(Cross-sectional) | SLE: N=18, 43.4 (12.0) years, 100% F<br>HC: N=20, 45.0 (11.9) years, 100% F | 38.1 (12.6) | SLEDAI: 5.2 (2.6)<br>SDI: 0.9 (1.2) | 100% - 18 SLE (cm) |
| Zimmerman, 2017, Brazil<br>(Cross-sectional) | SLE w/o CD: N=20, 43.0 (10.4) years, 100% F<br>SLE w/ CD: N=20, 42.6 (11.1) years, 100% F<br>HC: NR | 15.8 (10.0)<br>13.7 (10.5) | SDI: 1.5 (1.2)<br>SDI: 1.4 (1.7) | NR |
| Wiseman, 2017, UK<br>(Cross-sectional) | SLE: N=51, 48.8 (14.3) years, 92% F<br>HC: N=51, 44.9 (11.1) years, 76% F | Median=4.2<br>(IQR=2.0-12.3) | NR | 35% - 18/51 SLE (cr) |

| **First Author,**<br>**Year, Country**<br>**(Study Design)** | **Group: Size N, Age (years): Mean (SD) (or Range,**<br>**95% CI), Sex: % Females - F** | **Disease Duration**<br>**(years): Mean (SD)**<br>**(or Median, IQR)** | **Disease Activity (SLEDAI,**<br>**SLAM) & Damage (SDI): Mean**<br>**(SD)** | **Steroid Use: %**<br>**Patients**<br>**(cr - current,**<br>**cm - cumulative)** |
| --- | --- | --- | --- | --- |
| Cannerfelt, 2018,<br>Sweden<br>(Cross-sectional) | Non-NPSLE: N=27, 37.0 (9.4) years, 100% F<br>NPSLE: N=43, 39.4 (9.1) years, 100% F<br>HC: N=25, 39.6 (9.4) years, 100% F | 10.5 (7.8) | SLEDAI: 2.0 (2.6), SDI: 0.5 (0.9)<br>SLEDAI: 2.5 (3.5), SDI: 0.8 (1.2) | 90% - 63/70 SLE<br>(cr) |
| Corrêa, 2018,<br>Brazil<br>(Cross-sectional) | SLE w/o mem deficit: N=47, 45.0 (10.2) years, 97.9% F<br>SLE w/ mem deficit: N=20, 39.4 (11.4) years, 90% F<br>HC: N=22, 44.5 (8.8) years, 81.8% F | 14.4 (3.6)<br>15.3 (5.1) | SDI: 1.5 (1.2)<br>SDI: 1.4 (1.7) | Not used for 0.5<br>year pre-MRI (cr) |
| Nystedt, 2018,<br>Sweden<br>(Cross-sectional) | Non-NPSLE: N=25, 36.0 (CI=32.4–39.7) years, 100% F<br>NPSLE: N=39, 37.5 (CI=34.6–40.3) years, 100% F<br>HC: N=20, 37.3 (CI=33.1–41.5) years, 100% F | 11.2<br>(CI=8.1–14.4)<br>11.8<br>(CI=9.1–14.5) | SLEDAI: 2.0 (CI=0.9–3.1),<br>SDI:0.5 (CI=0.1–0.9)<br>SLEDAI: 2.7 (CI=1.5–3.8), SDI:<br>0.8 (CI=0.4–1.2) | 76% - 9/25 Non-NPSLE (cr)<br>87% - 34/39<br>NPSLE (cr) |
| Wiseman, 2018,<br>UK<br>(Cross-sectional) | SLE: N=47, 48.5 (13.7) years, 91.5% F<br>HC: NR | Median=4.0<br>(IQR=2.0–9.8) | NR | 36% - 17/47 SLE<br>(cr) |
| MacKay, 2019,<br>USA<br>(Longitudinal) | SLE: N=20, 41.0 (10.3) years, 90% F<br>*Follow-up (14.9 months):* N=13<br>HC: N=14, 42.0 (10.7) years, 85.7% F | 13.8 (9.2) | SLEDAI: 2.3 (2.1), SDI: 0.9 (1.1)<br>*Follow-up:* SLEDAI: 3.0 (2.8),<br>SDI: 1.4 (1.6) | 84% - 17/20 SLE<br>(cr) |
| DiFrancesco,<br>2020, USA<br>(Cross-sectional) | cSLE: N=11, 19.5 (4.2) years, 91% F<br>HC: N=11, 17.0 (6.8) years, 91% F | 6.3 (4.3) | SLEDAI: 5.7 (4.7), SDI:<br>Median=0 (Range=0–4) | 82% - 9/11 cSLE<br>(cr) |
| Mårtensson,<br>2021, Sweden<br>(Cross-sectional) | SLE: N=69, 35.8 (Range=18–51) years, 100% F<br>HC: N=24, 36.8 (Range=23–52) years, 100% F | NR | NR | NR |
| Qian, 2022,<br>Singapore<br>(Cross-sectional) | SLE: N=20, 36.1 (10.6) years, 80% F<br>HC: N=61, 29.2 (9.4) years, 48% F | 7.2 (5.3) | SLEDAI: 3.7 (3.5), SDI>0 in 5/20<br>patients | 95% - 19/20 SLE<br>(cr & cm) |
| Julio, 2023,<br>Brazil<br>(Cross-sectional) | cSLE: N=71, 24.6 (4.6) years, 90% F<br>aSLE: N=49, 33.2 (3.7) years, 90% F<br>HC: N=58, 29.9 (3.6) years, 72% F | 11.8 (4.8)<br>11.3 (4.1) | NR | 78% - 55/71 cSLE<br>(cr)<br>86% - 4/49 aSLE<br>(cr) |
| aSLE=Adult-onset SLE; CD=Cognitive Dysfunction; CI=Confidence interval; cSLE=Childhood-onset SLE; EM=Episodic memory; HC=Healthy controls; IQR=Interquartile range; MS=Multiple sclerosis; NR=Not available; NPSLE=Neuropsychiatric SLE; SD=Standard deviation; SDI=Systemic Lupus International Collaborating Clinics–ACR Damage Index; SLAM=Systemic Lupus Activity Measure; SLEDAI=SLE Disease Activity Index. |  |  |  |  |

The number of patients with clinical NPSLE diagnosis was reported in 16 cohorts and it was quite variable (ranged 0-100% of their respective cohorts), with half of these cohorts describing proportions over 50%, and three studies mentioning CD as a previous clinical NPSLE symptom. SDI ranged from 0-2.3 and it was the most frequently reported clinical variable (12/18 studies). Nine articles reported SLEDAI ranging from 2.0-15.3, whereas one manuscript utilized the Systemic Lupus Activity Measure (SLAM=9.9 ± 4.9). Disease duration was stated in 16/18 papers, and spanned 0.5-38 years since SLE symptom onset. Additionally, information regarding glucocorticoid exposure, retrieved from either current or cumulative use, was provided in 14 papers, and in 10 of them over 75% of patients were exposed to some form of glucocorticoids.

### Neurocognitive Assessments

Studies assessed cognitive performance of SLE patients with diverse approaches classified into the following categories: neuropsychological batteries, computerized batteries, screening tests, and incomplete/mixed designs^38^. A full neurocognitive characterization of the reviewed SLE cohorts is available as Supplementary Table S1.

Comprehensive neuropsychological batteries comprised at least four tests sensitive to specific cognitive domains (≥ 2 assessed domains) that have been validated in SLE (including the ACR-SLE, and the modified version endorsed by the Childhood Arthritis & Rheumatology Research Alliance – CARRA)^38^. These included, for example: Wechsler Intelligence Scales (Adult – WAIS-3 and WAIS-4, Children – WISC-4, and Abbreviated – WASI in 7/18 studies, Continuous Performance Test (CPT – simple attention, 8/18), Stroop Color-Word Test (SCWT – attention and working memory, 5/18), California Verbal Learning Test (CVLT) and Rey Auditory Verbal Learning Test (RAVLT) – verbal memory and learning, 3/18 each, Trail Making Test (TMT – visual-spatial processing, psychomotor speed and memory, 3/18), Hayling Test (HT – executive skills, specifically response initiation and inhibition, 3/18).

Six studies included neuropsychological batteries^23,24,27,28,30,36^. While the above standard neuropsychological batteries were still the most frequently employed category, they require lengthy assessments that must be administered by clinical psychologists^8^. Thus, computerized batteries, such as CNS Vital Signs (3/18) and Automated Neuropsychological Assessment Metrics – ANAM (2/18) have become more popular in SLE and they were utilized in 5 manuscripts^19,21,25,29,33^. These computerized batteries are shorter than traditional neuropsychological batteries, have demonstrated high sensitivity and specificity with the ACR-SLE battery, and can be administered by less clinically specialized staff^38,39^.

Screening tests are recurrently used to evaluate multiple domains and provide a quick global estimate of cognitive function. These screening tools were applied in six studies: Mini Mental State Examination (MMSE, 6/18)^20,30–32,34,35^, Montreal Cognitive Assessment (MoCA, 3/18)^20,26,34^, Addenbrooke’s Cognitive Examination (ACE, 3/18)^20,32,34^, and National Adult Reading Test (NART, 2/18)^20,34^. The use of screening tests has increased during the last decade as they might complement subjective assessments and guide additional neuropsychological tests to evaluate specific cognitive domains of interest in patients with SLE. Mixed/incomplete designs that included a combination of these screening tests and/or less than four domain-specific neuropsychological tests were reported in three papers^12,31,35^.

Lacking a generally accepted definition of CD in SLE, cut-offs for related deficits were selected in seven studies by choosing standardized z-scores −1.0 to −2.0 SD below the normative mean, either in at least two individual cognitive domains (if cut-off was lower than −1SD)^27,28^ or in only one domain (if cut-off was lower than −1.5 or −2 SD)^12,27,28,30,31,35,36^. Two of these seven studies exclusively focused on verbal memory (RAVLT)^12,30^.

### MRI Technical Details and Brain Structural Findings

Technical parameters, post-processing, and group differences in MRI metrics are available in Supplementary Table S2. In 10/18 studies brain MRI data was acquired with 3T scanners, while remaining studies were acquired at lower magnetic fields (2/18 at 2T and 6/18 at 1.5T). Regional volume was the most commonly evaluated structural MRI metric (8/9 studies) and it was mainly calculated from automated segmentations (4/8) and voxel-based morphometry (3/8). Both post-processing techniques employ semi-automated algorithms for volume quantification, reducing measurement susceptibility to operator skill. Additionally, segmentation methods (manual and automated) were targeted to specific brain regions in three studies (two in hippocampus, one in hippocampus and corpus callosum), therefore these studies did not evaluate potential abnormalities across the entire brain^29,32,35^. Diffusion MRI metrics from 9 DTI studies included FA (8/9), mean and directional diffusivities (MD, RD, AD – 4/9), as well as FA-weighted global and local brain structural connectivity metrics (1/9)^34^. They were mainly computed with tractography, tract-based spatial statistics, or voxel-wise analyses. One tractography study focused on the corpus callosum, cingulum and uncinate fasciculus tracts^19^ while another study solely retrieved metrics from corpus callosum automated segmentations^26^. IVIM-derived diffusion-perfusion metrics were examined in one study^27^.

#### Structural MRI

A summary of the structural MRI abnormalities in SLE reported across the studies is depicted in Figure 2, including T1-weighted MRI segmentation maps of GM and WM structures, and atlas-based parcellations of cortical structures. Lower total GM volume was reported in two SLE cohorts when compared to healthy controls. The hippocampus was the most frequently affected structure bilaterally, with smaller volumes reported in patients with SLE relative to controls (2/9)^32,35^, and worse hippocampus atrophy observed in patients with NPSLE (1/9)^29^ or CD (1/9)^36^. The next most frequent abnormalities were smaller frontal (2/9) and temporal (2/9) GM regions in adults and children with SLE and CD^28,31^. Additionally, regions within the frontal, temporal, and parietal cortices of SLE patients with memory deficits were thinner when compared to patients without memory deficits and controls^12^. Regarding longitudinal assessments, one study showed that the percentage of the SLE cohort with hippocampus atrophy increased by 23% when follow-up versus baseline MRI volumes were compared (from 47/107 patients with hippocampus atrophy at baseline to 40/60 patients at follow-up)^35^, while lower corpus callosum, frontal, dorsolateral, and medial temporal cortical volumes were reported in patients relative to controls during over a year follow-up period in another longitudinal study^31^.

**Figure 2:**
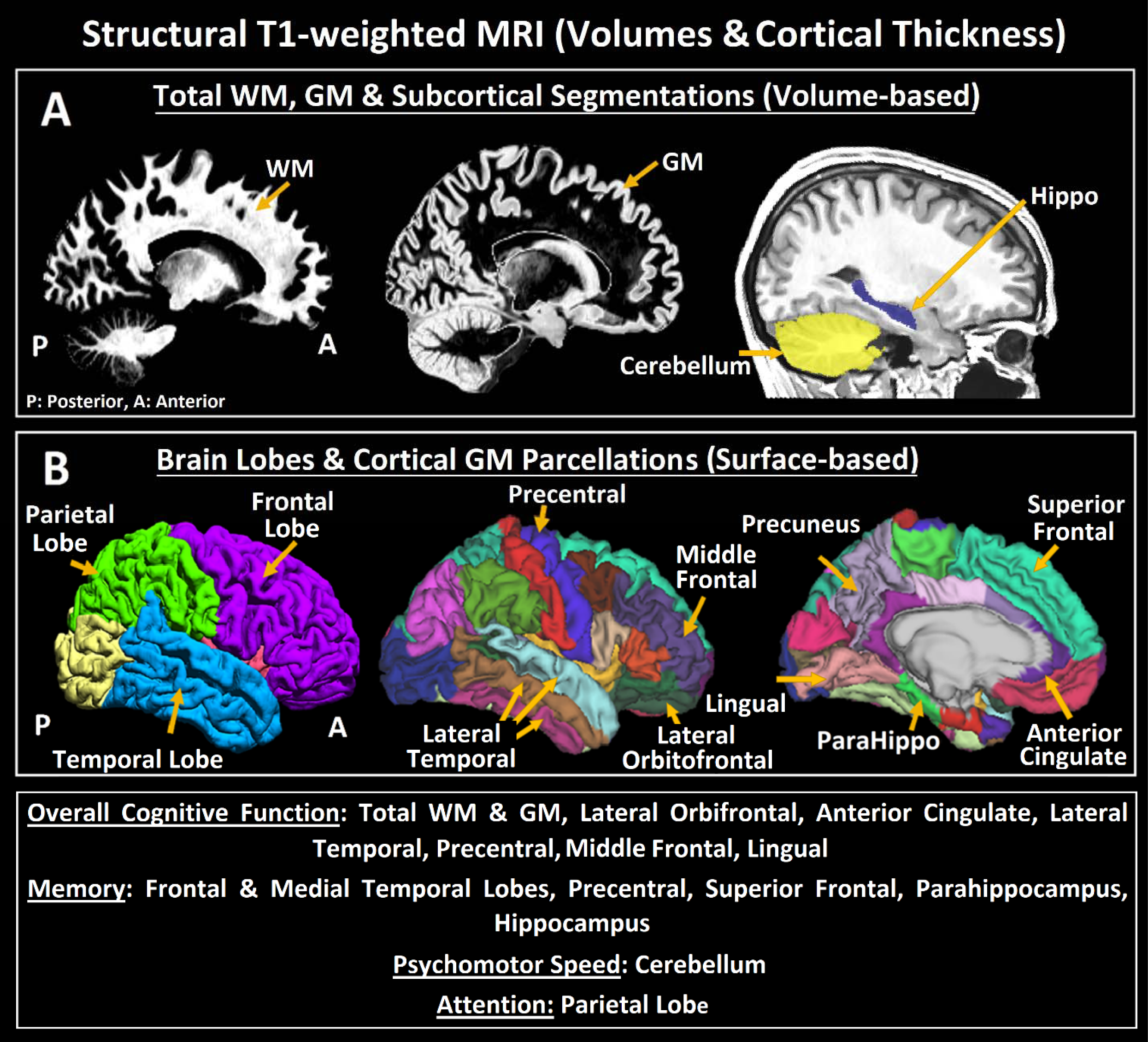
(A) Lower total white matter, grey matter, cerebellum, subcortical volumes, and (B) lower cortical volumes and thickness were commonly observed in patients with SLE (sagittal views). Structural MRI metrics were calculated from automatic segmentations and from parcellations labeled from a human brain atlas (Desikan-Killany). Atrophy was frequently observed in hippocampus and frontal cortex. Beyond general cognitive performance, memory, psychomotor speed and attention were the most frequently affected cognitive domains in SLE.

#### Diffusion MRI

A summary of the microstructural brain abnormalities on diffusion MRI in SLE reported across the studies in depicted in Figure 3, including WM atlas-based segmentations superimposed in a color-encoded FA map and an example of a 3D-rendered tract. The corpus callosum was the WM pathway most frequently damaged in SLE (6 studies), with lower FA (5/6), higher MD and RD (2/6), and higher AD and free-water (1/6) in patients with SLE when compared to controls^19,20,23,25,26,30^. Other frequently affected WM pathways were: the cingulum^19–21,25^ and fascicles connecting the frontal cortex in four studies (inferior fronto-occipital fasciculus, inferior and superior longitudinal fasciculus, uncinate fasciculus)^21,23,25,30^, and projection tracts in three studies (thalamic radiation, internal and external capsule, corticospinal tract, corona radiata)^20,23,30^

**Figure 3:**
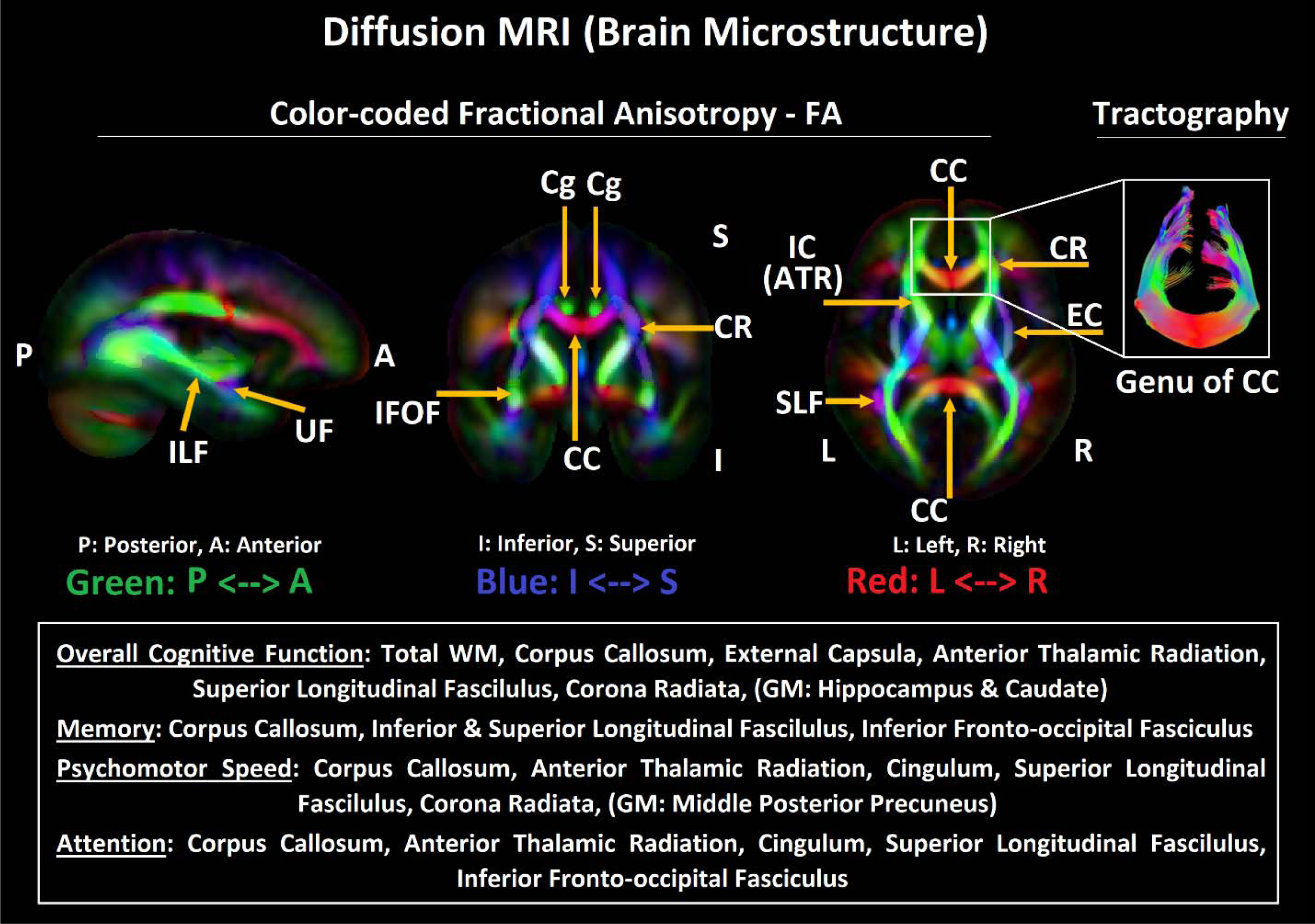
Abnormal widespread white matter microstructure (lower FA, higher diffusivities) wa reported in patients with SLE (sagittal, coronal and axial view), with a predilection for corpus callosum and periventricular/frontal tracts. Diffusion MRI metrics were mostly calculated with tractography (3D-rendered genu of the corpus callosum viewed from top) or from WM tract labels with tract-based spatial statistics. Beyond general cognitive performance, memory, psychomotor speed and attention were the most frequently affected cognitive domains in SLE.

One study evaluated diffusion metrics in GM and reported abnormalities in patients with cSLE, mainly higher D and D* in the precuneus/cuneus, occipital, post cingulate, and parietal regions^27^. Within SLE subgroups, one study reported large affected areas in SLE patients with and without memory deficits vs controls (lower FA and higher MD/RD)^30^. Another study evaluated patients with cSLE vs aSLE and reported lower FA and higher diffusivities in the corpus callosum in the former relative to the latter subgroup^26^, while no diffusion differences were observed between NPSLE and non-NPSLE patients in two manuscripts^19,24^. No changes in diffusion metrics in patients with SLE after the follow-up period were observed in the DTI publication with longitudinal data^25^.

### Cognitive Function and Relationships between Brain Structural Metrics

Poor overall cognition function was the most frequently reported metric of CD (8 studies). The cognitive domains most consistently impaired in SLE were: memory (visual, verbal, working, episodic, composite) in five articles^12,24,25,31,35^, followed by psychomotor speed and attention (simple, complex, sustained) in four^19,24,27,33^ and three studies^21,24,31^, respectively. One longitudinal study reported that the prevalence of CD after the follow-up period remained the same ^25^, while it increased by 34% (5 patients) after the follow-up period in another longitudinal study^35^. Brain structures/regions implicated in these cognitive domains are summarized in table 2, and their anatomic locations are shown in Figures 2 and 3.

**Table 2:**
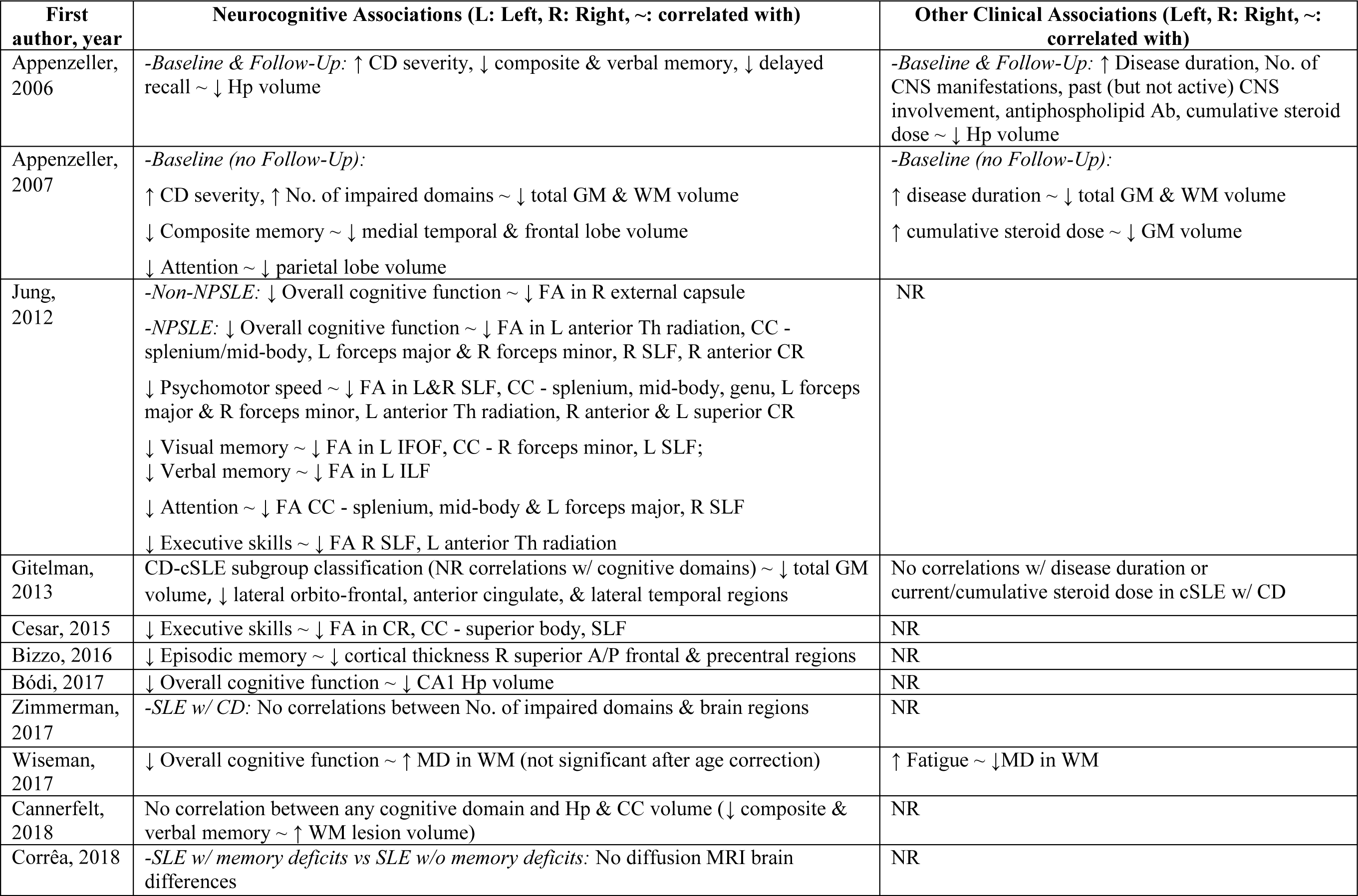
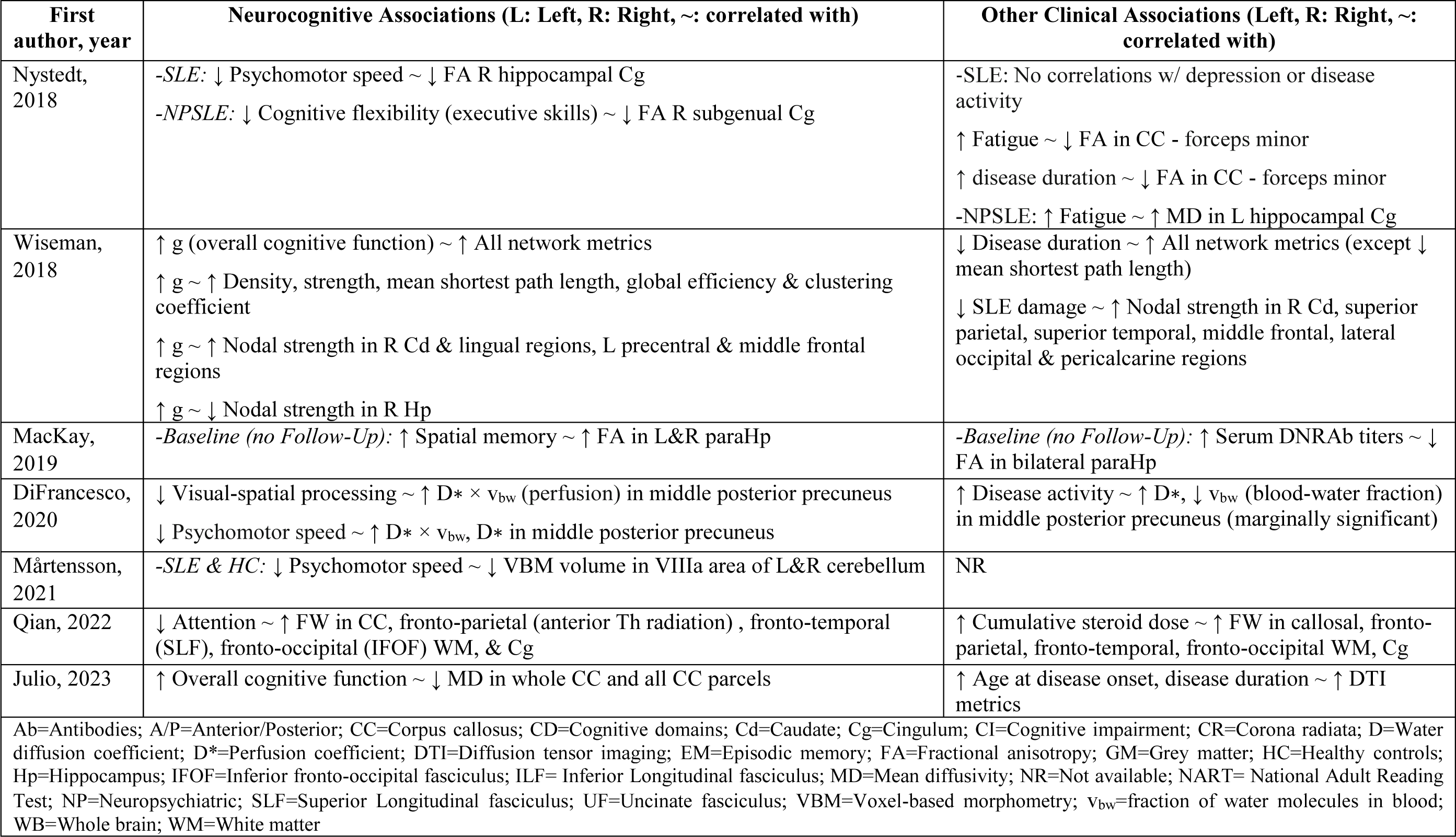
Associations between Brain Structural MRI Metrics, Cognitive Performance and Other Clinical Features in SLE Cohorts of Previous Studies.

With regard to structural brain MRI metrics, lower GM and WM volumes were associated with worse overall cognitive function and a greater number of impaired cognitive domains in patients with SLE^31^. Smaller volumes in the temporal and frontal lobes were associated with worse composite memory, while smaller volumes in the parietal lobe correlated with worse attention^31^. Abnormal hippocampus metrics frequently related to CD (3 studies), with smaller volumes linked with worse overall cognitive function^32,35^, memory (composite and verbal) and recall^35^. Other cortical and subcortical areas showing associations with CD were: frontal and precentral cortex (lower thickness with lower episodic memory), and the cerebellum (lower volumes and slower psychomotor speed)^12,33^.

With regard to diffusion MRI metrics, for total WM, higher MD was associated with lower overall cognitive function^20^. The corpus callosum was the WM structure that was most recurrently associated with CD (3/18 studies), with higher MD^26^ and lower FA^24^ correlating with worse overall cognitive function, lower FA^24^ and higher free water^21^ being associated with poor attention, and lower FA with visual memory and psychomotor speed^24^. Lower FA in the cingulum associated with slower psychomotor speed and worse cognitive flexibility (executive functioning domain)^19^, while higher free water was associated with worse attention^21^. Lower FA in anterior portions of the corona radiata (2/18), thalamic radiation (1/18), and right external capsula (1/18) correlated with lower overall cognitive function^24^ and executive skills^23^, with anterior thalamic radiation also correlating with worse processing speed^24^. Lower FA in the superior longitudinal fasciculus (2/18) was associated with lower overall cognitive function, visual memory, psychomotor speed, attention^24^ and executive skills^23^. Lower FA in the inferior fronto-occipital and longitudinal fasciculus respectively correlated with lower visual and verbal memory in one study^24^.

Additionally lower parahippocampal FA related to worse spatial memory^25^, higher node strength in the frontal cortex and in caudal/lingual regions respectively correlated to greater overall cognitive function and lower episodic memory^32^, and higher IVIM-derived perfusion in the precuneus associated with slower psychomotor speed and worse visual-spatial processing^25^.

### Associations between Clinical Variables and Abnormal Brain Structure (n=10) and CD (n=1)

Longer disease duration was related to lower total GM and WM volumes^31^, weaker network connectivity metrics in the whole brain^34^, and lower FA in the corpus callosum^19^. Greater SLE damage was associated with lower nodal strength in caudate and in several cortical regions in all brain lobes^34^, higher disease activity with higher water diffusion and lower blood-water fraction in precuneus^27^, higher levels of DNRAb serum titers with lower FA in parahippocampal areas^25^, and greater number of CNS manifestations with lower hippocampus volume^35^ (Table 2). Cumulative glucocorticoid dose was linked to lower GM volumes^31^ and higher free water in the corpus callosum, cingulum, and WM tracts connecting to the frontal cortex^21^. Greater fatigue was linked to higher MD in total WM^20^ and lower FA in the corpus callosum of patients with SLE, and to higher MD in the cingulum of patients with NPSLE^19^. One study reported worse overall cognitive function negatively associated with greater disease duration, higher expression of the inflammatory cytokine interleukin-6, and higher level of endothelial dysfunction antigens and activity (von Willebrand Factor)^20^.

## Discussion

To our knowledge, this is the first systematic review of neuroimaging literature on structural MRI abnormalities in SLE in relationship to CD. Our work included 18 peer-reviewed manuscripts and summarized their results in terms of clinical characterizations of SLE cohorts, neurocognitive assessments and CD definitions, structural brain MRI abnormalities and their links to CD and other related clinical factors in patients with SLE. We found that memory and attention, as well as psychomotor speed were the most consistently impaired cognitive domains in SLE when evaluated in relationship with structural brain alterations. CD in these domains correlated with abnormal MRI metrics (low volumes, abnormal microstructure), particularly in hippocampus and corpus callosum. Longer disease duration, higher cumulative glucocorticoid doses, and fatigue were disease factors often linked to regional brain structure abnormalities.

CD was associated with injury in several areas, with a particular emphasis in periventricular and frontal WM pathways (e.g., the corpus callosum), cortical frontal and parahippocampal regions, and certain subcortical GM structures (e.g., hippocampus) that are known to be involved in cognitive processes frequently affected in SLE patients. Lower volumes and cortical thinning were observed in fronto-temporal and hippocampal/parahippocampal regions in relationship to impairments in all cognitive domains in patients with SLE. Lower FA and higher diffusivities in WM tracts such as the corpus callosum and in subcortical structures, specifically the hippocampus, were utilized as indicators of microstructural brain alterations in NPSLE and non-NPSLE patients, and metrics in these regions were linked to poorer attention and impaired visual and working memory. Global and regional brain abnormalities in GM and WM were related to longer disease duration, NPSLE diagnosis, higher fatigue, greater disease activity, and higher cumulative glucocorticoid use, which suggests that brain damage, specifically in periventricular regions, could worsen with progressive pathology directly or indirectly caused by SLE. These results aligned with a longitudinal study that showed that hippocampus volumes, although affected early in SLE, further decreased with time and in relationship to factors such as greater total glucocorticoid dose, CD, and number of CNS manifestations^35^.

The hippocampus proper, and interconnected parahippocampal and periventricular regions are critical for memory and executive skills^40^. Neuronal injury in the hippocampus could extend to neighboring WM tracts as a consequence of anterograde or retrograde axonal degeneration and this mechanism has been proposed as a mediator of CD in SLE^7^ and MS^41^. Microstructural degeneration of these regions, could be also due to their preferential location in the brain, adjacent to cerebrospinal fluid and vascular spaces. It makes them particularly vulnerable to SLE pathology and treatment, including but not limited to microglial activation, as reported from in-vitro and mouse studies^42^ and glucocorticoid use^43^. Microstructural alterations in these regions could precede regional and global brain atrophy, and both microstructural and macro-structural abnormalities could even lead CD in SLE. However, to our knowledge, there have been only three MRI studies longitudinally evaluating structural brain metrics^10,31,35^, and one cross-sectional study that has combined both advanced structural and diffusion metrics in relationship to CD in SLE^23^. Additionally, only 2 studies evaluated CD in pediatric SLE populations^27,28^. Children with cSLE are at higher risk for developing CNS manifestations due to NPSLE, and they represent an opportunity to investigate the effects of SLE on the brain with little presence of comorbid conditions^44^. Studies of CD utilizing neuroimaging in cSLE may therefore provide particular insight into the mechanisms underlying the impact of SLE on the brain. In addition to structural brain abnormalities, factors that directly correlated to CD in patients with SLE included long disease duration and high expression of inflammatory cytokines. However, relationships between these factors and CD were only evaluated in one study^20^.

It is important to note that several methodological issues limited our interpretation of the findings. These include: (i) inadequately described demographic and disease related features in heterogeneous SLE cohorts; (ii) infrequent accounting of potential confounders such as glucocorticoid use, mood disorders, fatigue, disease activity, and duration; large variability in (iii) neuropsychological assessments utilized to evaluate CD and (iv) technical details in MRI scanners and acquisitions in studies in patients with SLE; (v) lack of harmonized neuroimaging analyses that combine structural MRI metrics from different modalities to evaluate both brain tissue morphology and microstructure; and (vi) lack of longitudinal data. These limitations indicate a need for consensus recommendations and guidelines regarding relevant demographic, clinical and cognitive function measures, and suitable technical MRI parameters and processing/post-processing pipelines. Such recommendations could inform collaborative neuroimaging studies in SLE across the globe, and create a knowledgebase of more comparable findings.

In conclusion, the results collected in this systematic review suggest that advanced structural MRI metrics can identify CNS abnormalities in patients with SLE and CD. Together with functional and metabolic neuroimaging tools, these metrics could serve as complementary diagnostic tools of NPSLE, as well as outcome measures in clinical trials focusing therapeutic interventions and neuroprotection and preserving cognitive function in SLE. However, improved characterization of SLE cohorts, guidelines for neuroimaging acquisitions and analyses, and more longitudinal studies are needed to further confirm the diagnostic and predictive ability of these metrics in SLE-related CD.

## Data Availability

All data produced in the present work are contained in the manuscript.

