## Supplementary Tables for "Effects of Systemic Lupus Erythematosus on the Brain: A Systematic Review of Structural MRI Findings and their Relationships with Cognitive Dysfunction"

**Table S1: Neurocognitive Characterization, Cognitive Domains, Neuropsychological Assessments, and Cut-offs for Cognitive Dysfunction (CD) Classification** **in SLE Cohorts**

| **First author, year** | **Neurocognitive Characterization (Clinical NPSLE Diagnosis & CD Status)** | **(Cognitive Domains)**  **Neuropsychological Assessments (Scoring)** | **CD Classification Cut-offs** |
| --- | --- | --- | --- |
| Appenzeller, 2006 | *Baseline:* 64/107 NPSLE, 30/64 – active CNS disease,  CD in 35/107 SLE (5 mild, 10 moderate, 20 severe CD)  *Follow-up:* 40/60 - active CNS disease,  CD in 40/60 SLE (6 mild, 12 moderate, 22 severe CD) | (Attention, executive skills, memory, visual-spatial processing, language, reasoning, psychomotor speed)  CVLT, MMSE, WAIS-4 (z-scores) | CD: z-score ≤ -2SD in any domain (CD: mild < 3, moderate 3-4, severe > 5 domains) |
| Appenzeller, 2007 | *Baseline:* 36/75 NPSLE, 15/36 – active CNS disease  *Follow-up:* NR | (Attention, executive skills, memory, visual-spatial processing, language, reasoning, psychomotor speed)  CVLT, MMSE, WAIS-4 (z-scores) | CD: z-score ≤ -2SD in any domain (CD: mild < 3, moderate 3-4, severe > 5 domains) |
| Jung, 2012 | -16/31 NPSLE  -NPSLE vs Non-NPSLE: ↓ psychomotor speed | (Overall cognitive function, attention, executive skills, psychomotor speed, memory)  COWAT, DS, HVLT, RCFT, TMT, WAIS-3, WRAT-3 (individual & total z-scores) | No CD cut-off |
| Gitelman, 2013 | -cSLE w/ CD: 6/8 NPSLE  -cSLE w/o CD: 1/14 NPSLE  -cSLE w/ CD vs cSLE w/o CD & HC: ↓ memory, psychomotor speed, visual-spatial processing, & overall cognitive function | (General cognition, attention, executive skills, psychomotor speed, memory, visual-spatial processing)  DKEFST, CPT-II, WASI, WISC-4, WRAML-2, Woodcock-Johnson –III Tests of Achievement | CD: z-scores ≤ -1SD in at least 2 domains or z-scores ≤ -2SD in at least one domain |
| Cesar, 2015 | -NPSLE: 6/23 – migraine, mood disorder & CD; 5/23 – migraine & CD; 2/23 – mood disorder & CD  -NPSLE vs HC: ↓ memory  -NPSLE vs MS: No differences in cognitive function | (Attention, executive skills, psychomotor speed, memory)  BVMT-R, CVLT, DKEFST, PASAT, SDMT (raw scores) | No CD cut-off |
| Bizzo, 2016 | -SLE w/o EM deficits: 14/34 NPSLE  -SLE w/ EM deficits: 10/17 NPSLE | (Memory – learning/recall)  RAVLT (individual & mean z-scores) | CD: Mean z-scores ≤ -1.5SD, or 2 or more individual z-scores ≤ -1.5SD |
| Bódi, 2017 | -SLE: No NPSLE diagnoses  -SLE vs HC: SLE patients showed a mild but significant CD on ACE scale | (Overall cognitive function)  MMSE, ACE (raw scores) | No CD cut-off |
| **First author, year** | **Neurocognitive Characterization (Clinical NPSLE Diagnosis & CD Status)** | **(Cognitive Domains)**  **Neuropsychological Assessments (Scoring)** | **CD Classification Cut-offs** |
| Zimmerman, 2017 | -SLE w/o CD: 11/20 NPSLE  -SLE w/ CD: 11/20 NPSLE, CD (15 mild, 4 moderate, 1 severe), ↑ CD in reasoning (57%), visual-spatial processing (38%), psychomotor speed (33%), executive skills (25%), language (24%) | (Attention, executive skills, language, reasoning, memory, visual-spatial processing)  BCT, BBNAB, HT, RAVLT, SCWT, TMT, verbal fluency tasks, WAIS-3, WCST (z-scores) | CD:  z-scores ≤ -2SD in any domain |
| Wiseman, 2017 | -SLE: 4/51 NPSLE, overall CD (MoCA – 19/50, ACER – 13/50, MMSE – 5/50 SLE)  -SLE vs HC: ↓ premorbid intelligence (NART) | (Overall cognitive function)  MoCA, ACER, MMSE, NART (raw scores) | No CD cut-off |
| Cannerfelt, 2018 | -SLE: 43/70 NPSLE (38/43 – migraine, 26/43 – CD, 25/43 – depression, 19/43 – anxiety, 18/43 – neuropathy) | (Memory, executive skills, psychomotor speed, attention, reasoning)  CNS-VS | No CD cut-off for group classification |
| Corrêa, 2018 | -SLE: NPSLE diagnosis – NR, no group differences in any domain except memory | (Overall cognitive function, attention, executive skills, psychomotor speed, memory, visual-spatial processing)  MMSE, RAVLT, BBNAB, WAIS-3, TMT, HT, SCWT (z-scores) | CD: RAVLT (mem) – total z-scores ≤ -1.5SD, or z-scores ≤ -1.5SD in ≥ 2 RAVLT domains |
| Nystedt, 2018 | -SLE: 39/54 NPSLE    -NPSLE vs Non-NPSLE: ↓ psychomotor speed, attention, cognitive flexibility (executive skills)  -NPSLE vs HC: ↓ all cognitive domains | (memory, executive skills, psychomotor speed, attention, reasoning)  CNS-VS | No CD cut-off |
| Wiseman, 2018 | -SLE: 3/47 active NPSLE, NART correlated w/ ↑ g (overall cognitive function) | (Overall cognitive function)  MoCA, ACER, MMSE, NART (raw scores) | No CD cut-off |
| MacKay, 2019 | -SLE-2 (*Baseline):* No NPSLE diagnoses, Match-to-Sample, Matching Grids, CPT scores ≤ -2SD in 3/20 (15%), 8/20 (40%), 1/20 (5%), respectively  -SLE-2 (*Follow-up):* ↑ Match-to-Sample & Matching Grids scores (still remained ≤ -2SD for same patients)  -SLE-2 vs HC: ↓ scores on all ANAM tests | (Attention, memory, visual-spatial processing)  Matching Grids, Match-to-Sample, CPT (ANAM battery throughput scores – combined RT & accuracy) | No CD cut-off |
| DiFrancesco, 2020 | -cSLE: No NPSLE or CD diagnoses | (Attention, psychomotor speed, memory, visual-spatial processing)  CPT-II, KABC, SEBC, WASI, WISC-4 (z-scores) | CD: z-scores ≤ -2SD in one domain, or z-scores ≤ -1SD in 2 domains |
| **First author, year** | **Neurocognitive Characterization (Clinical NPSLE Diagnosis & CD Status)** | **(Cognitive Domains)**  **Neuropsychological Assessments (Scoring)** | **CD Classification Cut-offs** |
| Mårtensson, 2021 | -SLE: 41/69 NPSLE | (memory, executive skills, psychomotor speed, attention, reasoning)  CNS-VS | No CD cut-off |
| Qian, 2022 | -SLE: NPSLE diagnosis – NR  -SLE vs HC: ↓ mean IQ, ↓ CPT | (Attention, memory, visual-spatial processing)  WAIS-4, ANAM battery (throughput scores) | No CD cut-off |
| Julio, 2023 | -cSLE: 53/71 NPSLE, 31/71 CD  -aSLE: 36/49 NPSLE, 16/49 CD | (Overall cognitive function)  MoCA | No CD cut-off |
| ACE=Addenbrooke’s Cognitive Examination; ACER=ACE-Revised; ANAM=Automated Neuropsychological Assessment Metric; BBNAB=Brazilian Brief Neuropsychological Assessment Battery NEUPSILIN; BCT= Bells Cancellation Test; BVMT-R=Brief Visuospatial Memory Test–Revised; CARRA= Childhood Arthritis & Rheumatology Research Alliance, CNS-VS= Central Nervous System Vital-Signs; COWAT=Controlled Oral Word Association Test; CPT= Continuous Performance Test; cSLE= Childhood-onset SLE; CVLT= California Verbal Learning Test; DKEFST=Delis-Kaplan Executive Function System Test; DS=Digit Symbol; HC=Healthy Controls; HT= Hayling Test; HVLT= Hopkins Verbal Learning Test; IQ=Intelligence Quotient; KABC= Kaufman Assessment Battery for Children; MMSE= Mini Mental State Examination; MoCA= Montreal Cognitive Assessment; NR=Not reported; NART= National Adult Reading Test; NPSLE= Neuropsychiatric SLE; PASAT= Paced Auditory Serial Addition Test; RAVLT= Rey Auditory Verbal Learning Test; RCFT= Rey Complex Figure Test; RT=Reaction Time; SEBC= Second Edition Block Counting; SCWT= Stroop Color-Word Test; SD=Standard Deviation; SDMT=Symbol Digit Modalities Test; TMT=Trail Making Test; WAIS-3= Wechsler Adult Intelligence-III; WAIS-4= Wechsler Adult Intelligence Scale–Revised; WASI=Wechsler Abbreviated Scale of Intelligence; WRAT-3=Wide Range Achievement Test–III, WCS=Wisconsin Card Sort; WISC-4=Wechsler Intelligence Scale for Children-IV; WRAML-2=Wide Range Assessment of Memory and Learning 2 | | | |

| **Table S2: MRI Protocol Acquisition Parameters, Post-Processing, and Neuroimaging Findings** | | | | |
| --- | --- | --- | --- | --- |
| **First author,**  **Year** | **Field Strength, Modality** | **Technical Parameters** | **Protocol Type; Post-Processing** | **Brain MRI Findings (L: Left, R: Right, ~: correlated with)** |
| Appenzeller,  2006 | 2T, Structural MRI | 0.8×0.8×3.0 mm | T1w-IR; manual Hp segmentation | -SLE vs HC: ↓ L&R Hp volume at baseline & follow-up  -SLE:   - Hp atrophy *(Baseline)*: 44% (47/107) – L Hp (20/47), R Hp (10/47), L&R Hp (17/47) - Hp atrophy *(Follow-up)*: 67% (40/60) – L Hp (4/40), R Hp (4/40), L&R Hp (32/40) |
| Appenzeller,  2007 | 2T, Structural MRI | 1.0×1.0×1.0 mm | T1w-GRE; VBM | -SLE vs HC: ↓ volume in CC, frontal, occipital, temporal & limbic lobes, cerebellum  -SLE:   - ↓ volume in CC in NPSLE w/ past vs active CNS involvement - ↓ WM volume in A/P CC, ↓ GM volume in frontal, dorsolateral & medial temporal regions *(Follow-up vs Baseline)* |
| Jung ^a^,  2012 | 1.5T, Diffusion MRI | 2 mm thick slices, 2 b_0_, 12 directions | DTI; TBSS | Group brain differences: NR |
| Gitelman, 2013 | 3T, Structural MRI | 1.0×1.0×1.0 mm | T1w-MPRAGE; VBM | -cSLE vs HC: No differences  -cSLE w/ CD vs cSLE w/o CD & HC: ↓ total GM volume, ↓ lateral  frontal, orbito-frontal, anterior cingulate, & lateral temporal areas |
| Cesar,  2015 | 3T, Structural & Diffusion MRI | 0.9x0.9x1.5 mm  1.1×1.1×3.0 mm, b=1000 s/mm^2^, 39 directions | 3D T1w-GRE; automatic volume segmentations    DTI; TBSS (Free-water corrected) | - NPSLE vs HC: ↑ MD/AD/RD in total WM  - NPSLE vs MS:   - ↑ WB, WM, & GM volume - ↓ FA in CC, ILF, IFOF, forceps major & minor, Cg, Th radiation |
| Bizzo,  2016 | 1.5T, Structural MRI | 1.3×1.0×1.3 mm | T1w-MPRAGE; surface-based analysis | -SLE w/ EM deficits vs HC: ↓ thickness L supra-marginal & superior temporal cortex  -SLE w/ EM deficits vs SLE w/o EM deficits: ↓ thickness R superior frontal, A/P middle frontal & precentral cortex |
| Bódi, 2017 | 3T, Structural MRI | 1.0×1.0×0.9 mm | T1w-MPRAGE; automatic volume segmentations, Hp subfields volume | SLE vs HC: ↓ volume CA1 & CA4-DG Hp subfields |
| **First author,**  **Year** | **Field Strength, Modality** | **Technical Parameters** | **Protocol Type; Post-Processing** | **Brain MRI Findings (L: Left, R: Right, ~: correlated with)** |
| Zimmerman, 2017 | 1.5T, Structural MRI | 1.3×1.0×1.3 mm | T1w-MPRAGE; automatic volume segmentations | -SLE w/ CD vs SLE w/o CD: ↓ L&R Hp and L Ag |
| Wiseman,  2017 | 1.5T, Diffusion MRI | 1.9×1.9×2.5 mm, b=1000 s/mm^2^, 32 directions | DTI (SS-EPI); tractography | -SLE vs HC: ↑ MD in all WM tracts, ↑ FA in CC - genu & L CST, ↓ FA in CC – splenium, L&R Cg |
| Cannerfelt, 2018 | 3T; Structural MRI | 1.0×1.0×1.0 mm | T1w-MPRAGE; automatic Hp & CC volume segmentations | NPSLE vs non-NPSLE: ↓ L&R Hp volume in NPSLE |
| Corrêa,  2018 | 1.5T, Diffusion MRI | 2.1×2.1×2.1 mm, b=900 s/mm^2^, 30 directions | DTI; TBSS | -SLE w/ memory deficits vs HC: ↓ FA, ↑ MD, RD in CC, L&R ILF, IFOF, SLF, UF, CST, cerebral peduncle, posterior Th radiation, external capsule, A-P limb internal capsule, anterior CR, R superior CR  -SLE w/o memory deficits vs HC: ↓ FA, ↑ MD, RD in same structures except R posterior limb internal capsule & R anterior CR |
| Nystedt, 2018 | 3T; Diffusion MRI | 2.0×2.0×2.0 mm, b=1000 s/mm^2^, 64 directions | DTI; CC, Cg, & UF tractography | -SLE vs HC: ↓ FA in R rostral Cg, CC mid-sagittal & forceps minor; ↑ MD in L hippocampal Cg  -Non-NPSLE vs HC: ↓ FA in R rostral Cg, CC - forceps minor; ↑ MD in L hippocampal Cg  -NPSLE vs HC: ↓ FA in CC – mid-sagittal & forceps minor; ↑ MD in L hippocampal Cg  -Non-NPSLE vs NPSLE: No differences |
| Wiseman,  2018 | 1.5T, Diffusion MRI | 1.9×1.9×2.5 mm, b=1000 s/mm^2^, 32 directions | DTI; tractography | -17 Node Network Hubs:   - ↑ Mean shortest path length, ↓ global efficiency, clustering coefficient & mean edge weight ~ ↑ age - ↑ Mean shortest path length, ↓ other metrics ~ ↑ WM ‘lesion’ volume |
| MacKay,  2019 | 3T, Diffusion MRI | 1.9×1.9×2.5 mm,  b=800 s/mm^2^, 5 b_0_, 33 directions | DTI (SS-EPI); voxel-wise analysis & tractography | -SLE-2 vs HC: ↓ FA & ↓ no. of streamlines in SLF, UF (insular cortex), hippocampal Cg, ILF, IFOF & CC - splenium  -SLE-2: No FA changes *(Follow-up vs Baseline)* |
| DiFrancesco, 2020 | 3T, Diffusion MRI | 3.8×3.8×5.0 mm, b=0-1000 s/mm^2^ (15 b-values), 3 $\perp$directions | DWI (EPI) w/ CSF suppression; voxel-wise IVIM | -cSLE vs HC:   - ↑ D∗ (D in blood) & D∗ × v_bw_ (perfusion) in cuneus, precuneus, superior occipital & posterior cingulate regions; ↑ D in precuneus & R inferior parietal regions - ↓ v_bw_ (blood-water fraction) in cuneus, precuneus, calcarine & middle-occipital regions; all abnormal metrics in middle posterior precuneus |
| **First author,**  **Year** | **Field Strength, Modality** | **Technical Parameters** | **Protocol Type; Post-Processing** | **Brain MRI Findings (L: Left, R: Right, ~: correlated with)** |
| Mårtensson,  2021 | 3T, Structural MRI | 1.0×1.0×1.0 mm | T1w-MPRAGE; VBM | -SLE vs HC: ↓ VBM volume in VIIIa and VIIa area of L&R cerebellum    -NPSLE vs non-NPSLE: No differences |
| Qian, 2022 | 3T, Diffusion MRI | 2.3×2.3×2.3 mm, b=1000 s/mm^2^, 8 b_0_, 61 directions | DTI (SS-EPI); TBSS & tractography (Free-water corrected) | - SLE vs HC:   - ↑ FW in CC, fronto-parietal, fronto-temporal, fronto-occipital WM, Cg - No FA, RD, AD differences (Free-water corrected); no WM SC differences - ↓ FA, ↑ RD & AD in WM (Not free-water corrected) |
| Julio, 2023 | 3T, Diffusion MRI | 2.0×2.0×2.0 mm, b=1000 s/mm^2^, 32 directions | DWI (DTI); automatic segmentation of CC & shape analysis | -SLE vs HC: ↓ mid-sagittal area in the post CC  -cSLE vs aSLE: ↓ FA, ↑ MD, ↑ RD, ↑ AD in whole CC and all but rostral CC parcels. |
| ⊥=Orthogonal; 3D=Three-dimensional; AD=Axial diffusivity; Ag=Amygdala; A/P=Anterior/Posterior; aSLE=Adult-onset SLE; CC=Corpus callosum; CI=Cognitive Dysfunction; CA=Cornu ammonis; Cg=Cingulum; CR=Corona radiata; CSF=Cerebrospinal fluid; cSLE=Childhood-onset SLE; D=Water diffusion coefficient; ; D*=Perfusion coefficient; DG=Dentate gyrus; DTI=Diffusion tensor imaging; EPI= Echo-planar imaging; EM=Episodic memory; FA=Fractional anisotropy; FOV=Field of view; FW=Free water; FWE= FW elimination; GM=Grey matter; GRE=Gradient echo; Hp=Hippocampus; IFOF=Inferior fronto occipital fasciculus; ILF= Inferior Longitudinal fasciculus; IR=Inversion recovery; IVIM=Intravoxel incoherent motion; MD=Mean diffusivity; MPRAGE=Magnetization Prepared Rapid Gradient Echo Imaging; MS=Multiple sclerosis; NPSLE=Neuropsychiatric SLE; RD=Radial diffusivity; SC=Structural connectivity; SLF=Superior Longitudinal fasciculus; SS-EPI=Single-shot EPI; T=Tesla; TBSS=Tract-based spatial statistics; Th=Thalamus; UF=Uncinate fasciculus; VBM=Voxel based morphometry; v_bw_=fraction of water molecules in blood; WB=Whole brain; WM=White matter  ^a^ No diffusion b-value or voxel size provided. | | | | |
